# Patients with Achilles Tendinopathy use compensation strategies to reduce tendon load during rehabilitation exercises

**DOI:** 10.1101/2024.04.26.24306424

**Authors:** Frea Deroost, Davide Petrella, Ine Mylle, Benedicte Vanwanseele

## Abstract

**Purpose:** This study aimed to determine differences in the Achilles tendon loading during rehabilitation exercises for Achilles tendinopathy (AT), and the ranking of these exercises in patients with AT and controls.

**Methods:** Sixteen patients with AT (5F & 11M, 44.1 ± 12.9yr) and sixteen controls (4F & 12M, 39.4 ± 15.6yr) performed rehabilitation exercises while 3D motion and ground reaction forces were measured. Musculoskeletal modeling was used to compute joint kinematics and triceps surae muscle forces. Individual triceps surae muscle forces were summed to estimate Achilles tendon load. Subsequently, peak Achilles tendon loading, loading impulse, loading rate, loading indexes (combining the different loading parameters), and ankle and knee angles at the time of peak loading were determined.

**Results:** Patients with AT have a significantly reduced peak loading of the Achilles tendon compared to controls during the exercises with the highest peak loading: unilateral heel drop with flexed knee (3.66 ± 0.90BW [AT] vs. 4.65 ± 1.10BW [Control], p=0.003, d=0.979) and walking (3.37 ± 0.49BW [AT] vs. 3.68 ± 0.33BW [Control], p=0.044, d=0.742). Furthermore, ankle dorsiflexion and knee flexion were reduced during unilateral heel drop with a flexed knee for the AT group. The ranking of exercises by peak loading or loading index was similar for people with and without AT. However, the ranking of exercises depends on the parameter used to define Achilles tendon loading.

**Conclusion:** During the highest load-imposing exercises, patients with AT employ compensatory strategies to reduce the load on their Achilles tendon. Clear instructions and feedback on the patient’s performance are crucial as altered exercise execution influences Achilles tendon loading.

## Introduction

The Achilles tendon is the largest and strongest tendon in the human body. It consists of three subtendons derived from the two heads of the gastrocnemius muscle, the gastrocnemius medialis (GM) and lateralis (GL), and the soleus muscle (SOL). However, due to its size and its functional requirements, the Achilles tendon is very susceptible to injury. Exposure to high or unusual loads can lead to Achilles tendinopathy (AT) (1), a frequently occurring overuse injury associated with impaired physical performance, pain, and swelling (2). The incidence of AT is 2.35 per 1000 registered persons per year in the general adult population (3), and its lifetime incidence can exceed 50% in male athletes (4).

Various treatments for AT are available with active treatments preferred over a wait-and-see approach (5). Triggering the Achilles tendon with appropriate load (i.e. an optimal mechanical trigger resulting in regeneration of the pathological tissue) plays an important role in the recovering process of AT as it results in improvement of patient symptoms, normalization of tendon structure and optimization of functional performance (9). Additionally, under- or overloading the tendon can have adverse outcomes, highlighting the importance of finding this sweet spot of optimal mechanical loading (10). When overloading the tendon, matrix and cell changes will further progress (e.g. increase in matrix disorganization and decreased collagen), while underloading causes a decrease in the mechanical integrity of the tendon (1). To provide this optimal stimulus to the tendon, exercise-based programs are commonly used during AT rehabilitation. Based on the original work of Curwin and Stanish (6) and Alfredson et al. (7), eccentric exercises that progressively increase in load are still considered the principal choice for conservative treatment of AT (8). However, about 24% to 45.5% of patients with AT do not benefit sufficiently from exercise-based rehabilitation (11), meaning that symptoms continue despite the treatment being followed. This implies that the rehabilitation of AT can still be improved. Nowadays, rehabilitation is based on the progression of pain (12). By adapting progression based on objective criteria of loading, better tendon remodelling can be pursued.

To better understand the load on the Achilles tendon during commonly used rehabilitation exercises, Baxter et al. (9) derived loading indexes for various exercises commonly used during AT rehabilitation in healthy participants. Using different parameters to quantify tendon load (i.e.: peak loading, loading impulse, loading rate), exercises were segregated into four tiers to gradually increase Achilles tendon loading during rehabilitation. Usually, peak loading is used as the main loading parameter because the magnitude of peak loading influences the amount of collagen production (9, 20). In addition, loading impulse is important as long-term and repetitive loading contributes to the development of AT (21). Loading rate is considered because both the presence of high forces and loading rates on the Achilles tendon may impede the healing process of AT, as these are linked to a reduction in the tendon’s mechanical and material properties (22). More recently, Devaprakash et al. (23) and Funaro et al. (24) have both developed an exercise progression based on the internal Achilles tendon strain during rehabilitation and functional exercises. Exposing the tendon to strains between five and six percent, which are associated with optimal anabolic tendon adaptation, may result in improved rehabilitation compared to exercise-based intervention where no specific strain is targeted (10). However, the loading indexes of Baxter et al. (9) and the Achilles tendon strains determined by Devaprakash et al. (23) and Funaro et al. (24) were examined in healthy participants. Whether those loading indexes and strains are similar for individuals with AT remains unknown.

Patients with Achilles tendinopathy experience pain, functional and structural changes that might influence the loading on the tendon during exercises. Initially, the pain is triggered by activities that increase stress on the tendon, and it can worsen as the pathology progresses, eventually interfering with daily activities (11). Pain can potentially make patients move differently, influencing loading during activities. In addition to pain, functional and structural differences have been found between patients with AT and controls. These differences include an increase in cross-sectional area of the Achilles tendon (13), with evidence of hypercellularity, collagen fibre disruption and increased ground substance (14, 15), and a lower Achilles tendon stiffness and Young’s modulus (13). The plantar flexor muscle also seems to be involved as weak ankle plantar flexor muscles were found to be a risk factor for developing AT. However, there is still a disagreement in the literature about whether there is a difference in maximal plantar flexor muscle strength in people with and without AT (16–19). As there might be an interaction between pain and force production and both these factors can influence movement execution and therefore the load applied on the tendon during rehabilitation exercises, it is uncertain if patients with AT perform rehabilitation exercises similarly to healthy individuals.

Previous research has already reported alterations in ankle and knee kinematics among individuals with AT during walking and running compared to healthy controls. Patients with AT tend to initiate walking and running with a more extended knee angle upon initial contact (25, 26). Additionally, during running, patients with AT ran with a reduced range of motion (ROM) at the knee joint when compared to healthy controls, leading to notable changes in muscle functioning (27). Nevertheless, it remains unclear whether kinematic alterations during rehabilitation exercises between healthy controls and patients with AT are present and if they influence the Achilles tendon load.

This study aims to determine whether the Achilles tendon load (considering peak Achilles tendon loading, loading impulse and loading rate) differed between patients with AT and healthy controls during typical rehabilitation exercises. Additionally, we aim to evaluate whether the ranking of different rehabilitation exercises, based on the peak loading and loading index, varied between patients with AT and healthy controls. By better understanding the load on the Achilles tendon, we aim to create a better rehabilitation strategy based on objective criteria of tendon load rather than the subjective feeling of pain. We hypothesize altered Achilles tendon loading in patients with AT compared to healthy controls during dynamic exercises.

## Materials and methods

### Participants

Data were collected from sixteen participants with AT and sixteen healthy control participants without pain in their Achilles tendon (Table 1). Before data acquisition started, participants were informed about the experimental protocol (approved by the local ethics committee KU/UZ Leuven (S63532)), screened for eligibility for the study by a physiotherapist and provided written informed consent. All participants were physically able to undertake the tests, had no lower limb surgery in the last ten years, had no pain in other places of the lower body and had no other suspected ankle pathologies. Participants with AT had symptoms for at least two months, with symptoms increasing during tasks such as running and jumping and had a local thickening at the midportion Achilles tendon diagnosed through ultrasound imaging by the physiotherapist during the screening. Healthy controls were matched with participants with AT based on their gender, age, height, body mass, type and amount of physical activity. The International Physical Activity Questionnaire (IPAQ), assessing physical activity and sedentary time, and the Victorian Institute of Sports Assessment-Achilles questionnaire (VISA-A), assessing the severity of the Achilles tendon pain, were administered to all participants upon inclusion.

**Table 1:** Anthropometric data and injury characteristics.

|  | **AT group (n=16)** | **Control group (n=16)** | **p-value** |
| --- | --- | --- | --- |
| Gender (Females/Males) | 5/11 | 4/12 | - |
| Age (yr) | 44.06 ± 12.92 | 39.44 ± 15.56 | 0.37 |
| Height (cm) | 1.80 ± 0.06 | 1.78 ± 0.09 | 0.46 |
| Body mass (kg) | 78.00 ± 13.94 | 76.63 ± 11.52 | 0.76 |
| Body Mass Index (kg/m<sup>2</sup>) | 23.89 ± 3.54 | 23.40 ± 4.81 | 0.82 |
| Physical activity (MET.min/wk) | 9357 ± 7779 | 7527 ± 5971 | 0.16 |
| VISA-A score | 57.28 ± 14.30 | 100 ± 0.00 | <0.001 |

\* Significant difference between groups with $p < 0.05$

### Protocol

Subjects warmed up by cycling for five minutes on a home trainer at a self-selected pace. Seven rehabilitation exercises based on the eccentric rehabilitation protocol of Alfredson et al. (7) were executed: bilateral heel rise, bilateral heel drop, bilateral heel drop with flexed knees, bilateral heel drop with an additional weight in each hand (2×3 kg), unilateral heel drop, unilateral heel rise, and unilateral heel drop with a flexed knee. First, the exercises were explained and demonstrated, after which the participants were given time to practice. Speed of the exercises was controlled by using a metronome set to 1 Hz to aid the participant to execute the full movement in three seconds. After the familiarization, the exercises were recorded and checked for speed and range of motion. Additionally, three functional exercises were performed: walking, squatting and lunging. All exercises were executed barefoot in random order per category (functional and rehabilitation exercises) and were repeated five times or more if there were multiple mistrials. Depending on the pain score experienced by the participant, as assessed using the numerical pain rating scale (30), exercises were performed only four (or fewer) times.

### Kinematics, kinetics and dynamic optimization

Motion capture data was collected using thirteen infrared motion-capture cameras (Vicon; Oxford Metrics, Oxford, United Kingdom, 100 Hz) that recorded three-dimensional motion of 49 retroreflective markers. Markers were placed according to an extended Plug-in-Gait marker model (31). Further, an AMTI force plate (embedded in the ground) with a sampling frequency of 1000 Hz (Advanced Mechanical Technology, Inc, Watertown, MA) was used to collect ground reaction force (GRF) data of the injured/matched leg. A low-pass filter of 6 Hz was used to filter GRF data using a 4th-order Butterworth filter. A threshold of 20 Hz was used to define the start of the stance phase during walking, squatting and lunges.

All markers were labelled in Vicon Nexus 2.12 (Oxford Metrics, Oxford, United Kingdom). A static labeled marker trial was used to scale the Gait2392-generic model in OpenSim 3.3 (OpenSim, Stanford, CA, USA) to match the anthropometry of each participant. The maximal isometric muscle forces of all muscles within the scaled musculoskeletal model were adjusted according to the participant’s body mass (32). Additionally, these forces were multiplied by a factor of 3 to prevent excessively elevated reserve actuator forces and to avoid excessive muscle activations. Thereafter, a Kalman smoothing algorithm was used to calculate the ankle, knee and hip joint angles (33). Next, a standard inverse dynamics approach in OpenSim was used to compute the ankle, knee and hip joint moments based on the output kinematic data, low-pass filtered at a frequency of 6 Hz, and the filtered GRF data. Further, a dynamic optimization approach was used to solve the muscle redundancy problem (34) to estimate the muscle forces.

The data was further analysed in MATLAB R2022b (The MathWorks Inc., Massachusetts, USA). For each exercise, three trials were selected for each participant based on the standard deviation of the maximal knee and ankle angle. Any trials with a standard deviation greater than 1.5 were excluded until three trials remained. Hereafter, we determined the start and end time of each trial and normalized the data. Data from walk and lunge exercises were splined based on the time at which the measured leg made ground contact. For the squat, hip height, based on the marker positioned at the left posterior superior iliac spine, was used to determine the start and end of the trial. Start and end of heel rise and heel drop exercises was determined based on the highest and lowest position of the heel marker. Subsequently, we calculated the Achilles tendon load during all exercises as the sum of the muscle forces of the SOL, GM and GL at every time point. Based on this Achilles tendon load during each exercise, peak loading, loading impulse and loading rate were calculated (9). The peak loading corresponds to the highest triceps surae force during each exercise. Loading impulse is the cumulative load on the tendon and was calculated by determining the area under the tendon load-time curve. Loading rate was calculated by determining the peak rate of loading over a 5% moving window of the exercise. Next, based on these three loading parameters, Achilles tendon loading indexes were calculated using the formula of Baxter et al. (9). First, we normalized each of these loading parameters by dividing the loading parameters for each exercise by the maximum value of the respective parameter across the different exercises and groups. Thereafter, the loading parameters were scaled using scaling factors of 0.5 for peak loading, 0.3 for loading impulse and 0.2 for loading rate. Loading indexes were subsequently derived by summing these normalized and scaled loading parameters. The loading indexes hence range from 0 to 1, where 0 corresponds to no load, and 1 indicates an exercise with the highest peak loading, loading rate and loading impulse among the tested exercises. Furthermore, the knee and ankle angle at the time of peak loading were determined. In addition, walking speed during ground contact was calculated based on the displacement in the direction of walking of the marker placed on the tenth thoracic vertebra. For the squat, four control participants were excluded from further analysis due to the failure of the dynamic optimization, caused by extremely high knee flexions (higher than 120 degrees), for which the utilized musculoskeletal model was not suited.

### Surface electromyography

Surface electromyography (EMG) (Zerowire, San Jose, CA, 1000 Hz) was used to measure the muscle activity of the GM, GL and SOL of the injured or matched leg. Before placing the EMG electrodes, the skin was shaved and cleaned with ethanol. Thereafter, the electrodes were placed following SENIAM-guidelines (36). Raw EMG signals were first band-pass filtered (20-400 Hz), then rectified and low-pass filtered at 6 Hz. Next, muscle activations were normalized to the muscle activation achieved during a maximal voluntary contraction of the specific muscle. The validation of the muscle activations from the musculoskeletal model was done by comparing the pattern of the experimental muscle activations with the modeled muscle activations during all rehabilitation and functional exercises for each participant.

### Statistics

Statistical analyses were conducted using the SPSS software (version 28.0.1.1 (v14); IBM Corp., Armonk, NY, USA). A Shapiro-Wilk test was used to test a normal distribution of the anthropometric data, the three loading parameters, and the ankle and knee angles at the time of peak loading. To compare anthropometric data, the different loading parameters, and the ankle and knee angles between the AT and control groups, different statistical tests were used depending on the normality of the distribution. For normally distributed loading parameters, Levene’s test was used to assess equal variances. If equal variances were observed, an unpaired T-test was performed, otherwise a Welch T-test was used. For non-normally distributed data, a Mann-Whitney U test was used. A significance level of p ≤ 0.05 was used. Furthermore, the effect sizes were quantified using Cohen’s d values and categorized as small, medium, and large for values of 0.2, 0.5, and 0.8, respectively. Additionally, Pearson correlation coefficients were computed to assess the linear relationship between peak Achilles tendon loading and ankle joint kinematics. The strength of the relationship between the parameters was categorized as small, moderate, strong and perfect for absolute Pearson correlation coefficients lower than 0.3, 0.7, 1 and equal to 1 respectively.

## Results

Based on the peak loading, unilateral heel drop with a flexed knee, followed by walking (Figure 1) elicited the highest load. During these exercises, a significantly lower peak loading in patients with AT was observed, more specifically during walking (3.37 ± 0.49BW [AT] vs. 3.68 ± 0.33BW [Control], p = 0.044, d = 0.742) and during unilateral heel drop with the knee flexed (3.66 ± 0.90BW [AT] vs. 4.65 ± 1.10BW [Control], p=0.003, d=0.979) (Figure 1). Other exercises showed trends and only small-to-medium effect sizes (all p > 0.090, d < 0.597) towards a lower Achilles tendon peak load in patients with AT compared to healthy controls. Overall, a similar ranking order of exercises was observed between healthy controls and patients with AT. However, in healthy controls, unilateral and bilateral heel drop ranked higher compared to the same exercises executed by patients with AT, while the lunge ranked lower (Figure 1).

**Figure 1:**
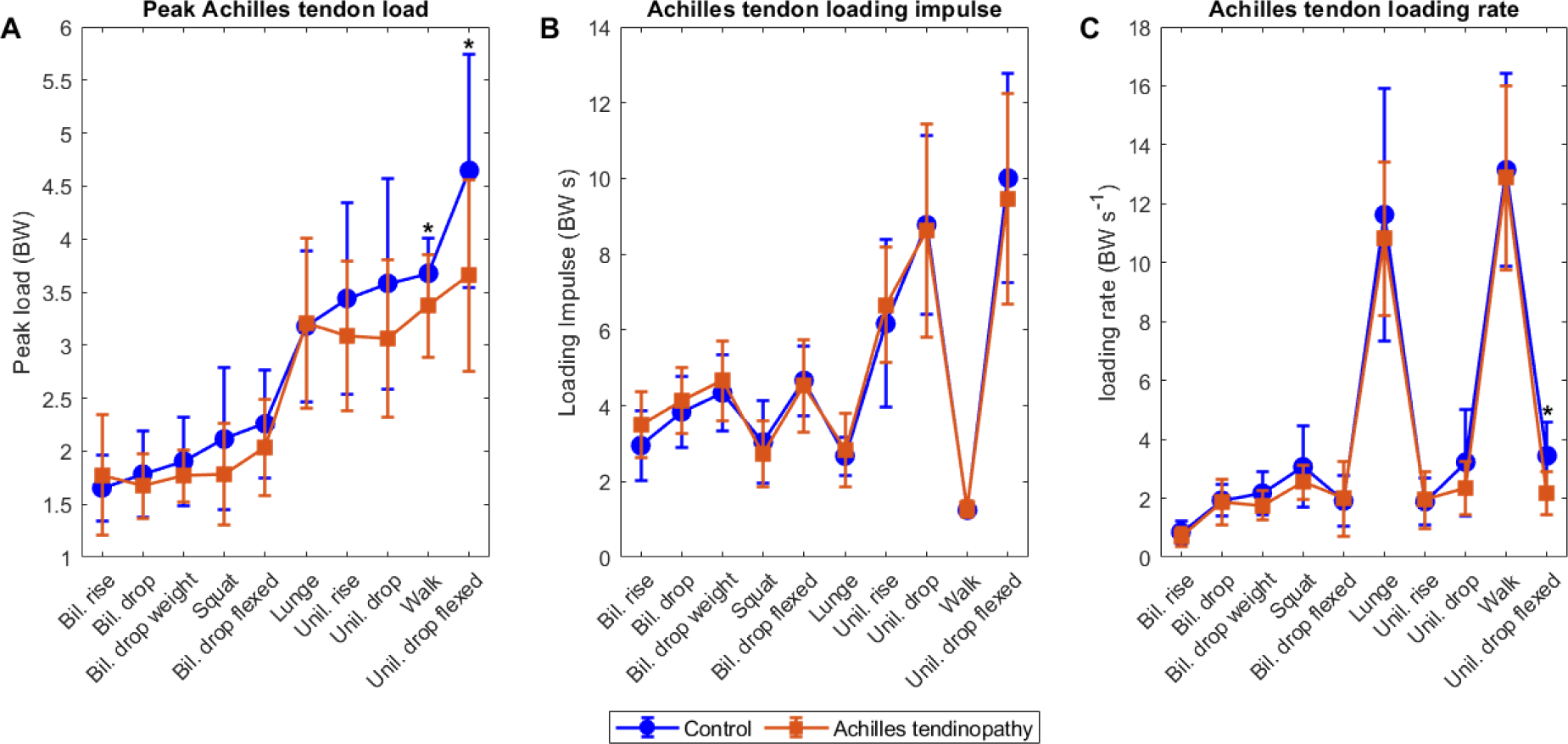
(A) Peak Achilles tendon loading (BW) of rehabilitation exercises for AT (orange) and control (blue) groups ranked from highest to lowest peak loading according to the control group. Significant differences (p < 0.05) between the AT and control group are marked with an asterisk. (B) Achilles tendon loading impulse (BW s) of rehabilitation exercises for AT (orange) and control (blue) groups ranked from highest to lowest peak loading according to the control group. Significant differences (p < 0.05) between the AT and control group are marked with an asterisk. (C) Achilles tendon loading rate (BW s^-1^) of rehabilitation exercises for AT (orange) and control (blue) groups ranked from highest to lowest peak loading according to the control group. Significant differences (p < 0.05) between the AT and control group are marked with an asterisk.

During unilateral heel drop with a flexed knee, patients with AT flexed the knee significantly less at the time of peak loading (p = 0.024, d = 0.838) and had a significantly lower ankle dorsiflexion angle (p = 0.045, d = 0.738) (Table 2). For all other exercises, a trend towards lower knee flexion and lower dorsiflexion at the time of peak loading was observed in patients with AT, except during the bilateral and unilateral heel raises.

**Table 2:** Mean knee and ankle angles at time of peak Achilles tendon load for AT patients and controls grouped by exercise. Exercises are ranked based on maximal peak loading of the Achilles tendon from figure 1. Positive angles correspond to knee flexion and ankle dorsiflexion, while negative angles indicate knee extension and ankle plantarflexion.

| Exercise | Knee angle (°) |  | Ankle angle (°) |  |
| --- | --- | --- | --- | --- |
|  | AT | Control | AT | Control |
| Unil. drop flexed | **51.07** ± **11.44** | **61.04** ± **12.33\*** | **28.71** ± **6.40** | **33.73** ± **7.17\*** |
| Walking | 8.29 ± 6.82 | 8.47 ± 3.78 | 12.63 ± 3.84 | 13.50 ± 3.61 |
| Lunge | 100.54 ± 10.63 | 102.45 ± 13.95 | 25.87 ± 7.26 | 25.82 ± 7.72 |
| Unil. rise | 7.22 ± 7.41 | 8.57 ± 8.24 | 13.20 ± 6.64 | 10.79 ± 6.04 |
| Unil. drop | 9.47 ± 8.64 | 10.54 ± 9.68 | 13.44 ± 5.72 | 16.55 ± 9.37 |
| Bil. drop flexed | 60.69 ± 12.61 | 68.62 ± 10.53 | 29.55 ± 6.83 | 32.52 ± 5.87 |
| Squat | 100.03 ± 12.63 | 100.24 ± 9.58 | 33.08 ± 5.48 | 33.68 ± 4.77 |
| Bil. drop weight | 6.60 ± 6.84 | 8.67 ± 6.34 | 11.29 ± 8.94 | 12.40 ± 12.28 |
| Bil. rise | 8.41 ± 6.53 | 7.02 ± 4.13 | 15.68 ± 9.94 | 11.33 ± 8.19 |
| Bil. drop | 6.07 ± 6.87 | 7.78 ± 6.52 | 12.45 ± 10.46 | 13.30 ± 11.18 |

\* Significant difference between groups with $p < 0.05$ .

In all exercises, an increase in ankle dorsiflexion was associated with an increase in Achilles tendon load. Strong associations were found during bilateral heel drop (r = 0.76, p < 0.001), bilateral heel drop with flexed knees (r = 0.70, p < 0.001), bilateral heel drop with additional weight (r = 0.73, p < 0.001) and squat (r = 0.74, p < 0.001).

Achilles tendon loading impulses during rehabilitation exercises were similar between individuals with and without AT (Figure 1). Remarkably, the highest Achilles tendon loading impulse occurred during unilateral heel drop with a flexed knee, while the lowest loading impulse was recorded during walking.

A significant lower loading rate was observed during unilateral heel drop with a flexed knee (2.16 ± 0.74BW [AT] vs. 3.45 ± 1.14BW [Control], p < 0.001, d = 1.338) (Figure 1) for the AT group. A similar trend for unilateral heel drop with extended knees (2.35 ± 0.91BW [AT] vs. 3.23 ± 1.81BW [Control], p = 0.187, d = 0.611) and bilateral heel drop with additional weight (1.76 ± 0.50BW [AT] vs. 2.18 ± 0.75BW [Control], p = 0.076, d = 0.659) was observed.

Based on the Achilles tendon loading indexes, the highest loading index was observed during the unilateral heel drop with a flexed knee in both groups (Table 3). In general, higher loading indexes were observed during unilateral exercises compared to bilateral exercises. Furthermore, higher values were observed during eccentric heel drop movements compared to the concentric heel rise counterpart (i.e. unilateral heel drop vs. unilateral heel rise and bilateral heel drop vs. bilateral heel rise). The ranking of the exercises from lowest to highest load based on the loading index was almost the same for patients with AT and healthy controls, only the squat was ranked higher in healthy controls.

**Table 3:** Loading index for functional and rehabilitation exercises in AT and control group based on the formula of Baxter et al. (25) with loading parameters normalized to the maxima from the captured exercises of both groups. With the ranking based on the loading index indicated between brackets, 1 corresponding to the lowest loading index and 10 to the highest loading index.

| Exercise | Loading index (ranking) |  |
| --- | --- | --- |
|  | AT | Control |
| Unilateral heel drop flexed | 0.71 (10) | 0.85 (10) |
| Unilateral heel drop | 0.62 (9) | 0.70 (9) |
| Walking | 0.60 (8) | 0.63 (8) |
| Lunge | 0.59 (7) | 0.60 (7) |
| Unilateral heel rise | 0.56 (6) | 0.58 (6) |
| Bilateral heel drop flexed | 0.38 (5) | 0.41 (5) |
| Bilateral heel drop weight | 0.36 (4) | 0.37 (4) |
| Bilateral heel drop | 0.33 (3) | 0.34 (2) |
| Squat | 0.31 (2) | 0.36 (3) |
| Bilateral heel rise | 0.31 (1) | 0.28 (1) |

## Discussion

This study demonstrated for the first time that peak loading is significantly lower in patients with AT in exercises with the highest load, i.e. unilateral heel drop with a flexed knee and walking. During these exercises, lower knee flexion and lower dorsiflexion at the time of peak loading was observed in patients with AT compared to healthy controls. In addition, the loading rate was also significantly lower in patients with AT during unilateral heel drop with a flexed knee. Despite these differences, the ranking of the exercises based on peak loading and loading indexes was similar between patients with AT and controls.

Patients with AT performed walking and unilateral heel drop with a flexed knee with significant lower peak loading in the Achilles tendon. The exercises are also the ones ranked highest based on peak loading and loading index. As the loading of the tendon is directly related to the plantar flexor muscle force, this reduced loading could be associated with weaker plantar flexors previously identified as one of the risk factors for AT (37). Furthermore, these findings are also in line with McAuliffe et al. (38) where patients with AT were found to have strength deficits in the symptomatic lower leg. In addition, patients with AT seem to adjust their movement during walking and unilateral heel drop with a flexed knee to execute the same exercise with a lower Achilles tendon load. These adjustments involve walking (non-significantly) slower (1.07 ± 0.17 m/s [AT] vs 1.15 ± 0.17 m/s [Control], p = 0.205) and exhibiting significantly less knee flexion and ankle dorsiflexion during the unilateral heel drop. However, it is difficult to determine whether the reduced Achilles tendon load is compensation for a lack of strength or a strategy to avoid pain. Pain-induced compensation potentially plays a role as patients with AT often experience heightened pain while performing exercises. Thereby, previous studies have reported that pain can also induce altered musculoskeletal activations (39–41). It is plausible that adjustments in movement execution due to pain among patients with AT played a role in the observed reduction in Achilles tendon loading between the two groups. Additionally, patients with AT exhibited a significantly lower loading rate during unilateral heel drop with a flexed knee compared to healthy individuals, which could also be a strategy to protect the tendon.

Our findings indicate that reduced knee flexion and ankle dorsiflexion at the time of peak loading are linked to a decrease in peak loading (and indirectly to loading index) among patients with AT. This confirms previous findings from Yeh et al. (28) who found that reaching more dorsiflexion during eccentric heel drop exercises resulted in higher peak Achilles tendon loads. The dorsiflexed ankle position places additional strain on the Achilles tendon as it elongates during the eccentric phase of the exercise, ultimately leading to greater mechanical loading. In addition, increasing the knee flexion will further lengthen the bi-articular gastrocnemii increasing the strain on the Achilles tendon. This observation aligns with our findings, demonstrating a positive correlation between greater dorsiflexion in the ankle and heightened peak loading of the Achilles tendon.

Additionally, diminished knee flexion during peak loading can change how the individual triceps surae muscles contribute to the overall triceps surae muscle force, which is also known as force-sharing within the triceps surae muscles. Alteration in force distribution may indicate a different loading pattern in the Achilles tendon of patients with AT compared to healthy individuals. Indeed, Mylle et al. (43) observed an altered load distribution in patients with AT during rehabilitation and walking exercises, which may potentially impact the sliding between the subtendons (44, 45). This observation suggests that the load distribution within the Achilles tendon could be an important factor in the development and progression of AT.

Although the current study is the first one to report Achilles tendon loading in patients with AT, comparing our healthy participants to previous literature, peak loading results for walking and variations in heel drop and heel rise exercises fall within the range previously observed (42). However, we did find lower peak loading for lunge and squat exercises, 3.2BW ± 0.7BW and 2.1BW ± 0.6BW respectively, compared to Baxter et al. (9) with peak loading of 2.1BW ± 0.6BW for lunge and 1.1BW ± 0.3BW for squat. The observed differences may stem from methodological disparities, such as the calculation of Achilles tendon load based on the plantar flexor moment using a fixed moment arm of 5cm. In contrast, in this study, Achilles tendon load was based on the sum of the forces exerted by the plantar flexor muscles, considering the changing moment arm of these muscles during the exercise. Moreover, the execution of exercises could also be slightly different. As evidenced by our results, variations in kinematics due to altered exercise execution may influence Achilles tendon loading.

Ranking the exercises from lowest to highest loading based on peak loading on the one hand and loading indexes on the other hand resulted in a similar ranking between patients with AT and healthy participants. However, when comparing the ranking based on peak loading and loading indexes clear differences in ranking are seen, which implies that considering different loading parameters has an influence on the overall Achilles tendon loading experienced. We have opted to employ the same scaling factors as suggested by Baxter et al. (9). However, altering the scaling factors for individual parameters could potentially impact the ranking of exercises. The relative importance attributed to peak loading, loading impulse, and loading rate may vary depending on the stage of rehabilitation. For instance, during the initial phases of rehabilitation, peak loading and loading impulse may be more important to increase the load-bearing capacity of the tendon (46). Conversely, in later stages, the clinician might opt to reassess and reorder exercises, focusing more on the rate at which the load is introduced to the tendon to facilitate sports re-entry and make it more functional.

Eccentric exercises that progressively increase in load are still considered the preferred treatment approach for AT (8). Our research indicates that the ranking of exercises is similar for healthy individuals and patients with AT. Moreover, our results highlight the importance of physiotherapists providing clear instructions and feedback about the performed exercises as small differences in knee and ankle angles can result in altered loading of the Achilles tendon. Additionally, our results and previous research (43) suggested that during daily activities such as walking, the Achilles tendon already undergoes a considerable amount of peak loading, yet with a lower loading rate. Therefore, it seems important to better account for this daily cumulative load during the rehabilitation. In addition, based on the IPAQ score, our participants with AT were already relatively active (Table 1) as nearly all participants with AT could be categorized as having high levels of physical activity (47). This suggests that the cumulative loading throughout the day was already high in our participants with AT, which may have contributed to the overload resulting in AT.

Results of this study should be interpreted in light of some limitations. Muscle forces are computed based on musculoskeletal modelling as direct measures are unavailable. To validate the model outcomes we compared the measured EMG signal with the estimated muscle activation patterns during the exercises (see Supplemental Figure 1, Supplemental Digital Content 1, example comparison EMG signal and modeled muscle activation). Similar patterns were observed during most activities. However, the modelled activation patterns were slightly different during unilateral heel drop with a flexed knee (SOL) and squatting (SOL) compared to the experimental muscle activities. The influence of pain caused by AT on exercise performance was not considered during the study. As a result, it is not clear whether the differences found in loading parameters are a result of the pain that the AT patients experience or due to a lack of muscle strength. Until now, there is no consensus about the relative importance of peak loading, loading impulse and loading rate. Therefore, the same scaling factors as stated in the article of Baxter et al. (9) were used to calculate the load indexes of the exercises. Lastly, we had to exclude four control participants for the squat due to the failure of the dynamic optimization caused by large knee flexions. This exclusion may have obscured differences between the AT and control groups, as the excluded control subjects performed the highest knee flexions, making the difference between the AT and control groups likely larger than currently observed.

## Conclusion

In conclusion, peak Achilles tendon load during rehabilitation exercises was lower in patients with AT compared to healthy controls during the exercises with the highest load. This discrepancy arises from patients with AT using compensatory strategies while performing these exercises, which reduces the load on the Achilles tendon. Good guidance from physiotherapists is crucial during rehabilitation, as even minor adjustments in exercise execution can exert a discernible impact on tendon loading. Furthermore, the ranking of exercises from lowest to highest Achilles tendon load was similar between patients with AT and healthy controls, suggesting that a progression of exercises based on the load measured in healthy subjects can be used for the rehabilitation of AT.

## Data Availability

All data produced in the present study are available upon reasonable request to the authors.

## Acknowledgment

The authors want to thank all the participants for their participation in the study. Funding for this research was provided by the FWO research project (G098222N).

## Conflict of Interest

The authors state that there is no conflict of interest. The results of the present study do not constitute endorsement by ACSM and are presented clearly, honestly, and without fabrication, falsification, or inappropriate data manipulation.

## Notes

### Competing Interest Statement

The authors have declared no competing interest.

### Author Declarations

The Ethical Committee of UZ/KU Leuven gave ethical approval for this work.

## References

1. Cook JL, Purdam CR. Is tendon pathology a continuum? A pathology model to explain the clinical presentation of load-induced tendinopathy. Br J Sports Med. 2009;43(6):409–16.

2. Maffulli N. Overuse tendon conditions: Time to change a confusing terminology. Arthroscopy: The Journal of Arthroscopic & Related Surgery. 1998;14(8):840–3.

3. de Jonge S, van den Berg C, de Vos RJ, et al. Incidence of midportion Achilles tendinopathy in the general population. Br J Sports Med. 2011;45(13):1026–8.

4. Kujala UM, Sarna S, Kaprio J. Cumulative Incidence of Achilles Tendon Rupture and Tendinopathy in Male Former Elite Athletes. Clinical Journal of Sport Medicine. 2005;15(3):133–5.

5. van der Vlist AC, Winters M, Weir A, et al. Which treatment is most effective for patients with Achilles tendinopathy? A living systematic review with network meta-analysis of 29 randomised controlled trials. Br J Sports Med. 2021;55(5):249–56.

6. Curwin S, Stanish W. Tendinitis: Its etiology and treatment. Collamore Press [Internet]. 1984;

7. Alfredson H, Pietilä T, Jonsson P, Lorentzon R. Heavy-Load Eccentric Calf Muscle Training For the Treatment of Chronic Achilles Tendinosis. Am J Sports Med. 1998;26(3):360–6.

8. Fares MY, Khachfe HH, Salhab HA, Zbib J, Fares Y, Fares J. Achilles tendinopathy: Exploring injury characteristics and current treatment modalities. The Foot. 2021;46:101715.

9. Baxter JR, Corrigan P, Hullfish TJ, O’Rourke P, Silbernagel KG. Exercise Progression to Incrementally Load the Achilles Tendon. Med Sci Sports Exerc. 2021;53(1):124–30.

10. Pizzolato C, Lloyd DG, Zheng MH, et al. Finding the sweet spot via personalised Achilles tendon training: the future is within reach. Br J Sports Med. 2019;53(1):11–2.

11. Maffulli N, Sharma P, Luscombe KL. Achilles Tendinopathy: Aetiology and Management. J R Soc Med. 2004;97(10):472–6.

12. Iversen JV, Bartels EM, Langberg H. The victorian institute of sports assessment - achilles questionnaire (visa-a) - a reliable tool for measuring achilles tendinopathy. Int J Sports Phys Ther. 2012;7(1):76–84.

13. Arya S, Kulig K. Tendinopathy alters mechanical and material properties of the Achilles tendon. J Appl Physiol. 2010;108(3):670–5.

14. Archambault JM, Wiley JP, Bray RC, Verhoef M, Wiseman DA, Elliott PD. Can sonography predict the outcome in patients with Achillodynia? Journal of Clinical Ultrasound. 1998;26(7):335–9.

15. Kader D, Saxena A, Movin T, Maffulli N. Achilles tendinopathy: some aspects of basic science and clinical management. Br J Sports Med. 2002;36(4):239–49.

16. Child S, Bryant AL, Clark RA, Crossley KM. Mechanical Properties of the Achilles Tendon Aponeurosis Are Altered in Athletes With Achilles Tendinopathy. Am J Sports Med. 2010;38(9):1885–93.

17. Nunes GS, Tessarin BM, Silva RS, Serrão FV. Relationship between the architecture and function of ankle plantar flexors with Achilles tendon morphology in ballet dancers. Hum Mov Sci. 2019;67:102494.

18. O’Neill S, Barry S, Watson P. Plantarflexor strength and endurance deficits associated with mid-portion Achilles tendinopathy: The role of soleus. Physical Therapy in Sport. 2019;37:69–76.

19. Crouzier M, Tucker K, Lacourpaille L, et al. Force-sharing within the Triceps Surae: An Achilles Heel in Achilles Tendinopathy. Med Sci Sports Exerc. 2020;52(5):1076– 87.

20. Arampatzis A, Karamanidis K, Albracht K. Adaptational responses of the human Achilles tendon by modulation of the applied cyclic strain magnitude. Journal of Experimental Biology. 2007;210(15):2743–53.

21. Wren TAL, Lindsey DP, Beaupré GS, Carter DR. Effects of Creep and Cyclic Loading on the Mechanical Properties and Failure of Human Achilles Tendons. Ann Biomed Eng. 2003;31(6):710–7.

22. Starbuck C, Bramah C, Herrington L, Jones R. The effect of speed on Achilles tendon forces and patellofemoral joint stresses in high-performing endurance runners. Scand J Med Sci Sports. 2021;31(8):1657–65.

23. Devaprakash D, Graham DF, Barrett RS, et al. Free Achilles tendon strain during selected rehabilitation, locomotor, jumping, and landing tasks. J Appl Physiol [Internet]. 2022; doi:10.1152/japplphysiol.00662.2021.

24. Funaro A, Shim V, Crouzier M, Mylle I, Vanwanseele B. Subject-Specific 3D Models to Investigate the Influence of Rehabilitation Exercises and the Twisted Structure on Achilles Tendon Strains. Front Bioeng Biotechnol [Internet]. 2022;10 doi:10.3389/fbioe.2022.914137.

25. Hein T, Janssen P, Wagner-Fritz U, Haupt G, Grau S. Prospective analysis of intrinsic and extrinsic risk factors on the development of <scp>A</scp> chilles tendon pain in runners. Scand J Med Sci Sports [Internet]. 2014;24(3) doi:10.1111/sms.12137.

26. Bramah C, Preece SJ, Gill N, Herrington L. Is There a Pathological Gait Associated With Common Soft Tissue Running Injuries? Am J Sports Med. 2018;46(12):3023–31.

27. Azevedo LB, Lambert MI, Vaughan CL, O’Connor CM, Schwellnus MP. Biomechanical variables associated with Achilles tendinopathy in runners. Br J Sports Med. 2009;43(4):288–92.

28. Yeh C, Calder JD, Antflick J, Bull AMJ, Kedgley AE. Maximum dorsiflexion increases Achilles tendon force during exercise for midportion Achilles tendinopathy. Scand J Med Sci Sports. 2021;31(8):1674–82.

29. Martin RL, Chimenti R, Cuddeford T, et al. Achilles Pain, Stiffness, and Muscle Power Deficits: Midportion Achilles Tendinopathy Revision 2018. Journal of Orthopaedic & Sports Physical Therapy. 2018;48(5):A1–38.

30. Jensen MP, McFarland CA. Increasing the reliability and validity of pain intensity measurement in chronic pain patients. Pain. 1993;55(2):195–203.

31. Vicon. Plug-in Gate Reference Guide. https://docs.vicon.com/display/Nexus29/Plug-in+Gait+Reference+Guide. 2016;

32. Handsfield GG, Meyer CH, Hart JM, Abel MF, Blemker SS. Relationships of 35 lower limb muscles to height and body mass quantified using MRI. J Biomech. 2014;47(3):631–8.

33. De Groote F, De Laet T, Jonkers I, De Schutter J. Kalman smoothing improves the estimation of joint kinematics and kinetics in marker-based human gait analysis. J Biomech. 2008;41(16):3390–8.

34. De Groote F, Pipeleers G, Jonkers I, et al. A physiology based inverse dynamic analysis of human gait: potential and perspectives. Comput Methods Biomech Biomed Engin. 2009;12(5):563–74.

35. Andersson JAE, Gillis J, Horn G, Rawlings JB, Diehl M. CasADi: A Software Framework for Nonlinear Optimization and Optimal Control. Math Program Comput. 2019;11(1):1–36.

36. Hermens HJ, Freriks B, Disselhorst-Klug C, Rau G. Development of recommendations for SEMG sensors and sensor placement procedures. Journal of Electromyography and Kinesiology. 2000;10(5):361–74.

37. Mahieu NN, Witvrouw E, Stevens V, Van Tiggelen D, Roget P. Intrinsic Risk Factors for the Development of Achilles Tendon Overuse Injury. Am J Sports Med. 2006;34(2):226–35.

38. McAuliffe S, Tabuena A, McCreesh K, et al. Altered Strength Profile in Achilles Tendinopathy: A Systematic Review and Meta-Analysis. J Athl Train. 2019;54(8):889–900.

39. Hodges PW, Tucker K. Moving differently in pain: A new theory to explain the adaptation to pain. Pain. 2011;152(3):S90–8.

40. Graven-Nielsen T, Arendt-Nielsen L. Peripheral and central sensitization in musculoskeletal pain disorders: An experimental approach. Curr Rheumatol Rep. 2002;4(4):313–21.

41. Merkle SL, Sluka KA, Frey-Law LA. The interaction between pain and movement. Journal of Hand Therapy. 2020;33(1):60–6.

42. Demangeot Y, Whiteley R, Gremeaux V, Degache F. The load borne by the Achilles tendon during exercise: A systematic review of normative values. Scand J Med Sci Sports. 2023;33(2):110–26.

43. Mylle I, Crouzier M, Hollville E, Bogaerts S, Vanwanseele B. Triceps surae muscle forces during dynamic exercises in patients with Achilles tendinopathy: A cross-sectional study. Scand J Med Sci Sports. 2023;33(11):2219–29.

44. Maas H, Finni T. Mechanical Coupling Between Muscle-Tendon Units Reduces Peak Stresses. Exerc Sport Sci Rev. 2018;46(1):26–33.

45. Handsfield GG, Inouye JM, Slane LC, Thelen DG, Miller GW, Blemker SS. A 3D model of the Achilles tendon to determine the mechanisms underlying nonuniform tendon displacements. J Biomech. 2017;51:17–25.

46. Scattone Silva R, Song KE, Hullfish TJ, Sprague A, Silbernagel KG, Baxter JR. Patellar Tendon Load Progression during Rehabilitation Exercises: Implications for the Treatment of Patellar Tendon Injuries. Med Sci Sports Exerc. 2024;56(3):545–52.

47. Craig CL, Marshall AL, Sjöström M, et al. International Physical Activity Questionnaire: 12-Country Reliability and Validity: Med Sci Sports Exerc. 2003;35(8):1381–95.

