## Supplemental Figure 1 for "Patients with Achilles Tendinopathy use compensation strategies to reduce tendon load during rehabilitation exercises"

Supplemental Figure 1: Example comparison EMG signal and modeled muscle activation for all exercises with modeled (solid line) and experimental (dashed line) for the three triceps surae muscles (with yellow = SOL, orange = GM and blue = GL) for one participant. The modeled muscle activations were multiplied by 2.5 (except lunge, bilateral heel drop with flexed knee and squatting which was multiplied by 4) to better compare the pattern with the experimental muscle activation.

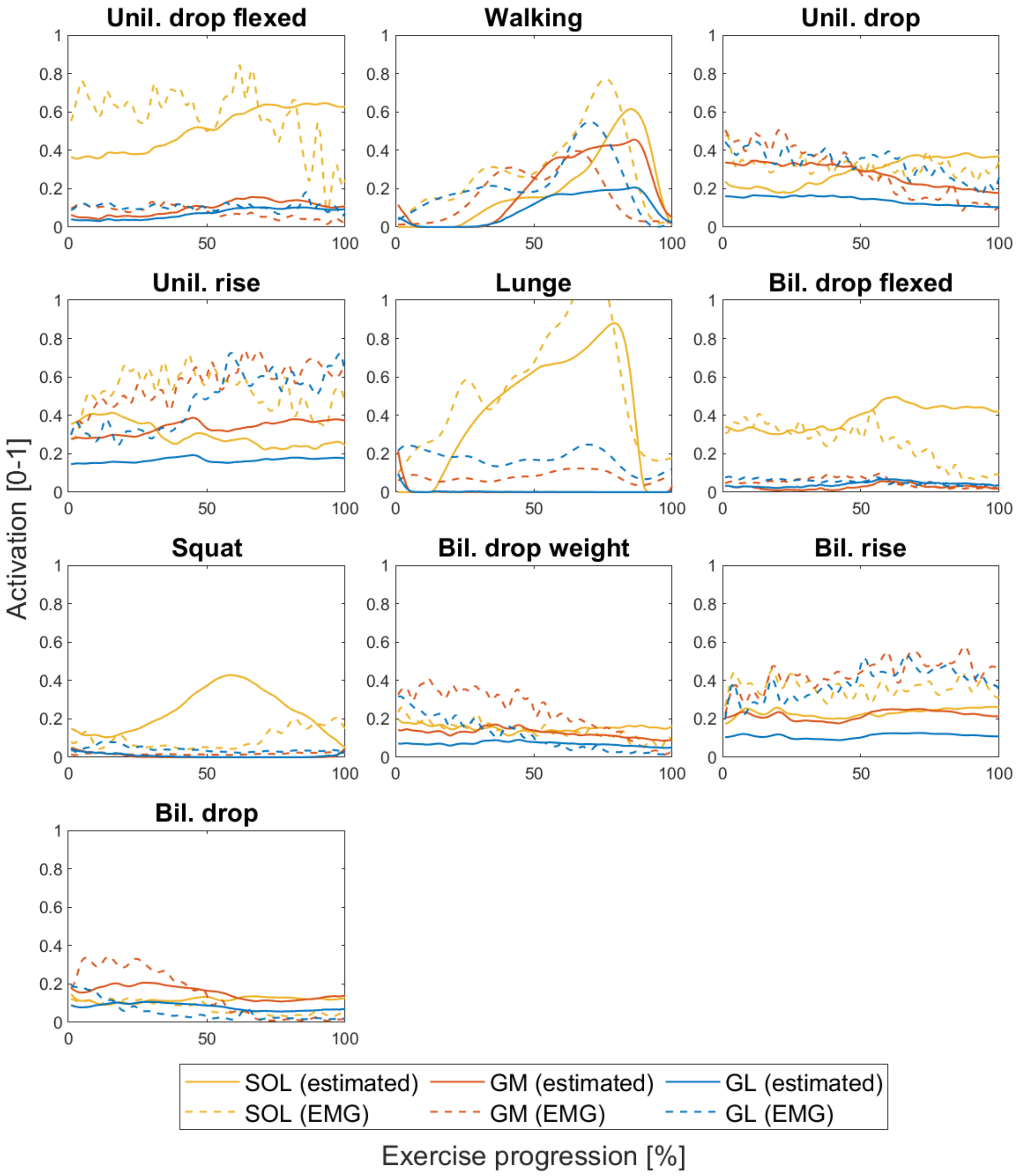
